# Population movement, city closure and spatial transmission of the 2019-nCoV infection in China

**DOI:** 10.1101/2020.02.04.20020339

**Authors:** Siqi Ai, Guanghu Zhu, Fei Tian, Huan Li, Yuan Gao, Yinglin Wu, Qiyong Liu, Hualiang Lin

## Abstract

The outbreak of pneumonia caused by a novel coronavirus (2019-nCoV) in Wuhan City of China obtained global concern, the population outflow from Wuhan has contributed to spatial expansion in other parts of China. We examined the effects of population outflow from Wuhan on the 2019-nCoV transmission in other provinces and cities of China, as well as the impacts of the city closure in Wuhan. We observed a significantly positive association between population movement and the number of cases. Further analysis revealed that if the city closure policy was implemented two days earlier, 1420 (95% CI: 1059, 1833) cases could be prevented, and if two days later, 1462 (95% CI: 1090, 1886) more cases would be possible. Our findings suggest that population movement might be one important trigger of the 2019-nCoV infection transmission in China, and the policy of city closure is effective to prevent the epidemic.

## Introduction

In December 2019, a series of pneumonia cases caused by a novel coronavirus, namely 2019-nCoV, emerged in Wuhan, the capital city of Hubei Province in China (1), Similar with severe acute respiratory syndrome (SARS), the Wuhan pneumonia outbreak was highly suspected to be linked to the wild animals in the seafood market, although the definitive source of this virus was not clear yet (2).

As of Jan 31, 2020, the infection has been transmitted to all the provinces in China and a few other countries. Epidemiology evidence shown that most of the cases had a history of living or travelling to Wuhan, the household cluster cases and cases of health-care workers indicated the human-to-human transmission route (3), which might be the reason for a rapid increasing rate of infection across the country and globally (4).

Considering the person-to-person transmission and the large travel volume during the traditional Chinese New Year (the largest annual population movement in the world), it is expected that the population movement would lead to further expansion of the infection, so the government imposed a lockdown on Wuhan City at 10:00 am on January 23, as well as some other cities later on. However, an estimated 5 million individuals had already left Wuhan for the holiday or travelling, some of which rushed out after the lockdown announcement (5). In addition, the novel coronavirus is infectious during the incubation period and when the symptoms are not obvious, which is likely to make the huge floating population potential sources of infection (6). Therefore, it is reasonable to hypothesize that the population transported from Wuhan may have a significant impact on the potential outbreaks in other parts of China.

Recent studies on the novel coronavirus pneumonia focused more on its etiology (7, 8), transmission route (9, 10), and epidemiological characteristics (11, 12), there is still a lack of investigating the relationship between the migrating population and the outbreak, which is of great importance for making intervention policies. Thus, we conducted this study with the following objectives: 1) to depict the impacts of the population movement on the spatial transmission of the 2019-nCoV cases at the provincial and city levels in China; 2) to estimate the potential outbreak risk at areas with the population outflowed from Wuhan; 3) to evaluate the effectiveness of the city closure measures on the epidemic control.

## Methods

### Data collection

The data on the daily number of cases were derived from the real-time update of the China Health Commission (http://www.nhc.gov.cn/), 2019-nCoV epidemic report on Phoenix and Dingxiangyuan website. The case definition has been introduced previously (13, 14). In brief, a confirmed case was defined as a pneumonia that was laboratory confirmed 2019-nCoV infection with related respiratory symptoms and a travel history to Wuhan or direct contact with patients from Wuhan. We collected each reported 2019-nCoV case in mainland China till 12:00 pm January 31, 2020.

### The floating population

As the city closure took place at 10:00 AM Jan 23^rd^, 2020, and the incubation period of the infection was estimated to be about 3-7 days (15), we obtained the daily index of population outflow from Wuhan and the proportion of the daily index from Wuhan to other provinces and top 50 cities, from January 1 to 31 in 2020, the information was retrieved through the Spring Festival travel information of China released by Baidu Qianxi. The data is consisted of Baidu Location Based Services (LBS) and Baidu Tianyan, using the positioning system and transportation information system, which be display dynamic visual regional population outflow in real-time. Data of Baidu Qianxi was freely available to the public (http://qianxi.baidu.com).

### Statistical analysis

#### Number of 2019-nCoV cases per unit outflow population

The daily index of population outflow from Wuhan to other provinces and top 50 cities was obtained by multiplying the daily index of population outflow within Wuhan by the corresponding proportion for each province. To evaluate the effect of prevention and control measures of the local government, we calculated the number of 2019-nCoV cases per unit outflow population, the formula can be specified as:

The total index of outflow population from Wuhan from Jan 1 to 31:

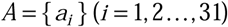

The daily proportion of the daily index from Wuhan to other provinces from Jan 1 to 31:

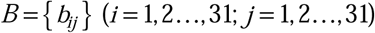

The cumulative 2019-nCoV cases in each province from Jan 1 to 31:

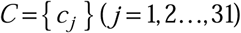

The total index of population inflow from Wuhan to other provinces:

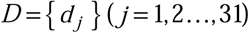

Number of cases per unit outflow population for each province:

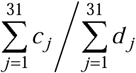

#### Evaluation of the effects of earlier city closure dates

After the city closure was taken in force, some population still outflowed from Wuhan. We subtracted the outflow index on Jan 23 from the average outflow index from Jan 24 to 31, to obtain the index of outflow population within Wuhan reduced by the advance city closure on Jan 23 and 22 (the advance outflow index). And then we calculated the average proportion of the outflow index from Jan 24 to 31 for each province (the average proportion). The number of cases reduced by the advance city closure in each province was estimated by multiplied the advance outflow index by the average proportion and one corresponding unit. The formula can be specified as:

The reduced index of outflow population:

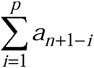

Here, n was the date that government announced city closure; p was the advance days of city closure.

The increased index of outflow population:

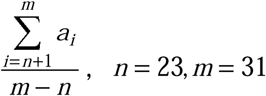

The net loss of index of outflow population:

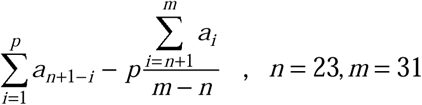

The average proportion of the outflow index from Wuhan into each province during the city closure:

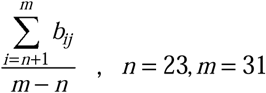

The net loss index of outflow population caused by advance Wuhan city closure for each province:

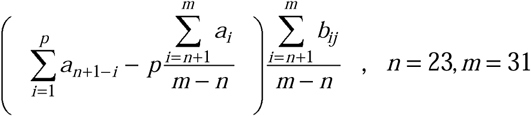

The total reduced number of 2019-nCoV cases:

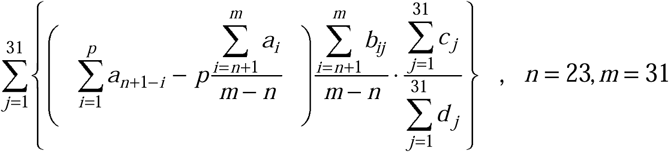

Similarly, we evaluated the impacts of one-day and two-day delayed city closure. We took the average index of the population outflow between Jan 21 and 23 as the daily index of population outflow before the city closure, and used the same calculation method to estimate the index of population outflow within Wuhan increased by the delayed city closure on Jan 24 and Jan 25 (the delayed outflow index). We multiplied the delayed outflow index by the average proportion and one corresponding unit to estimate the increased number of cases caused by one-day and two-day delayed city closure in each province.

## Results

As of January 31, 2020, a total of 11791 confirmed cases and 259 deaths due to the 2019-nCoV infection were reported in China, which were widely dispersed in all of 31 provincial administrative areas. The overall trend of 2019-nCoV cases is upward. During the period of January 11, 2020 through January 31, the cumulative number of cities infected with 2019-nCoV cases increased rapidly (appendix). A total of 313 cities in mainland China reported the occurrence of 2019-nCoV infection, of which the number of reported cases from January 20 to 29 increased rapidly from 7 to 313, an increase of nearly 44 times. Among the cities, 97 of them belong to the regions with high population exodus out of Wuhan, and 7138 cases have been reported, accounting for 83·23% of the total reported cases outside Wuhan (appendix).

Population outflow of Wuhan could be divided into four periods based on the migration data of Baidu (figure 1 and 2). In the first stage from Jan 1 to Jan10, migrant population flowed out of Wuhan normally and returned to hometown without the influence of the epidemic situation. At this point, the mean daily index of population outflow was 58039·71 (95% CI: 48883·38-66454·00), and 2019-nCoV cases were mainly distributed in Wuhan. From beginning of Spring Festival travel rush on Jan 10 and announcement of 2019-nCoV infection, the mean index of population outflow rose to 66777·98 (95% CI: 61125·90-72962·78), which increased by 15·1% compared with the previous period. Meanwhile, 2019-nCoV cases gradually appeared in Wuhan and several nearby cities. The third period was from January 20 to 23 when Academician Nanshan Zhong disclosed the human-to-human transmission and a large number of Wuhan residents fled from Wuhan with increasing panic. The corresponding mean index of population outflow of this period surged to 112385·88 (95% CI: 107367·44-118403·21), which increased by 93·6% comparing to the earlier period. Thereafter, a strict city closure policy was implemented to prohibit the population relocation from Wuhan. In the wake of sharp decline of outflow population from Wuhan, the index reduced to 9180·29 (95% CI: 3055·35-19101·40), falling by 91·8% comparing with the third period and the effective controlling of the outflow population prevented suspected cases from spreading to the whole country, however, 2019-nCoV cases has been widely dispersed in all of 31 provinces.

**Figure 1.**
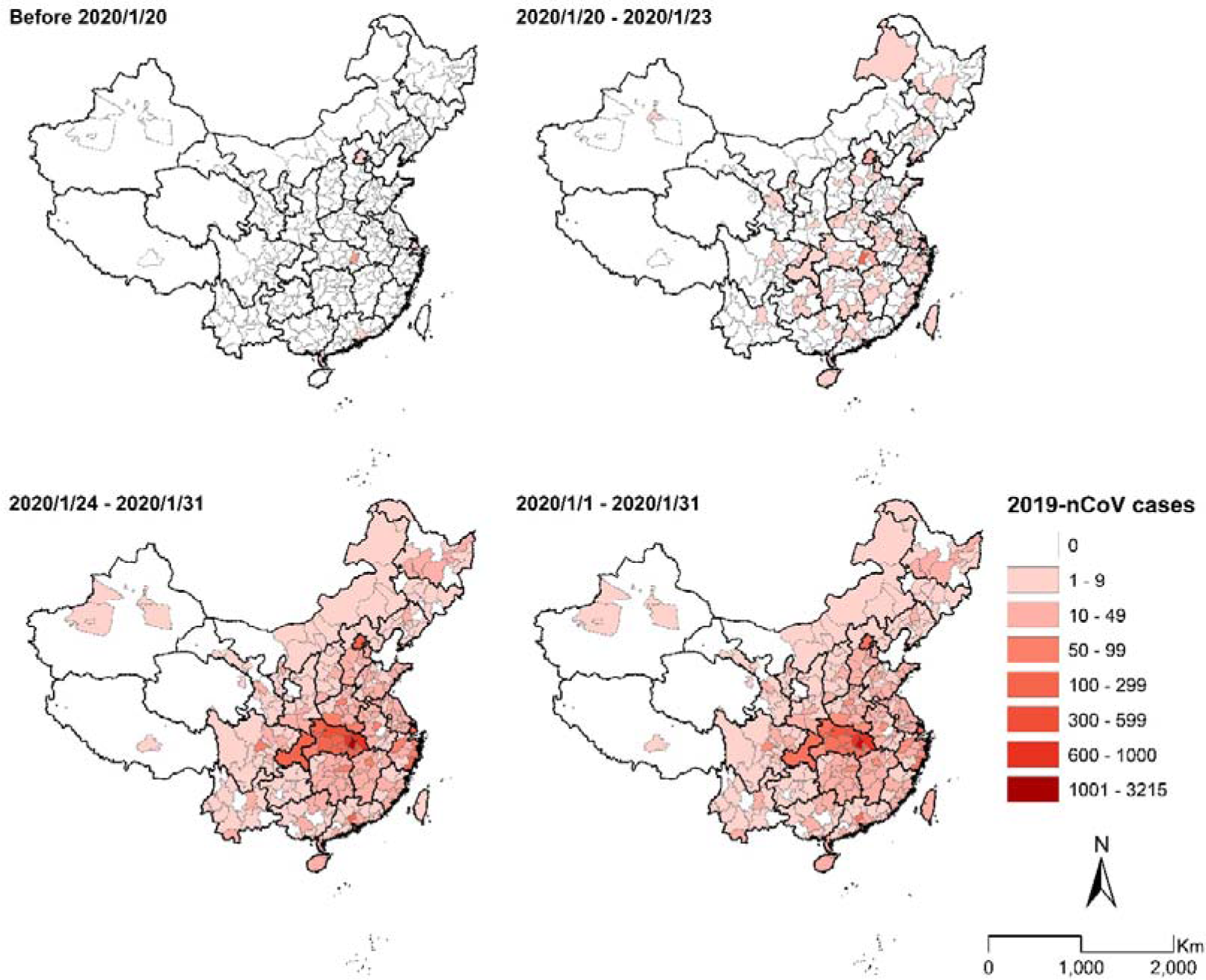
The spatial distribution of 2019-nCoV cases in China during period of January 1-31, 2020.

**Figure 2.**
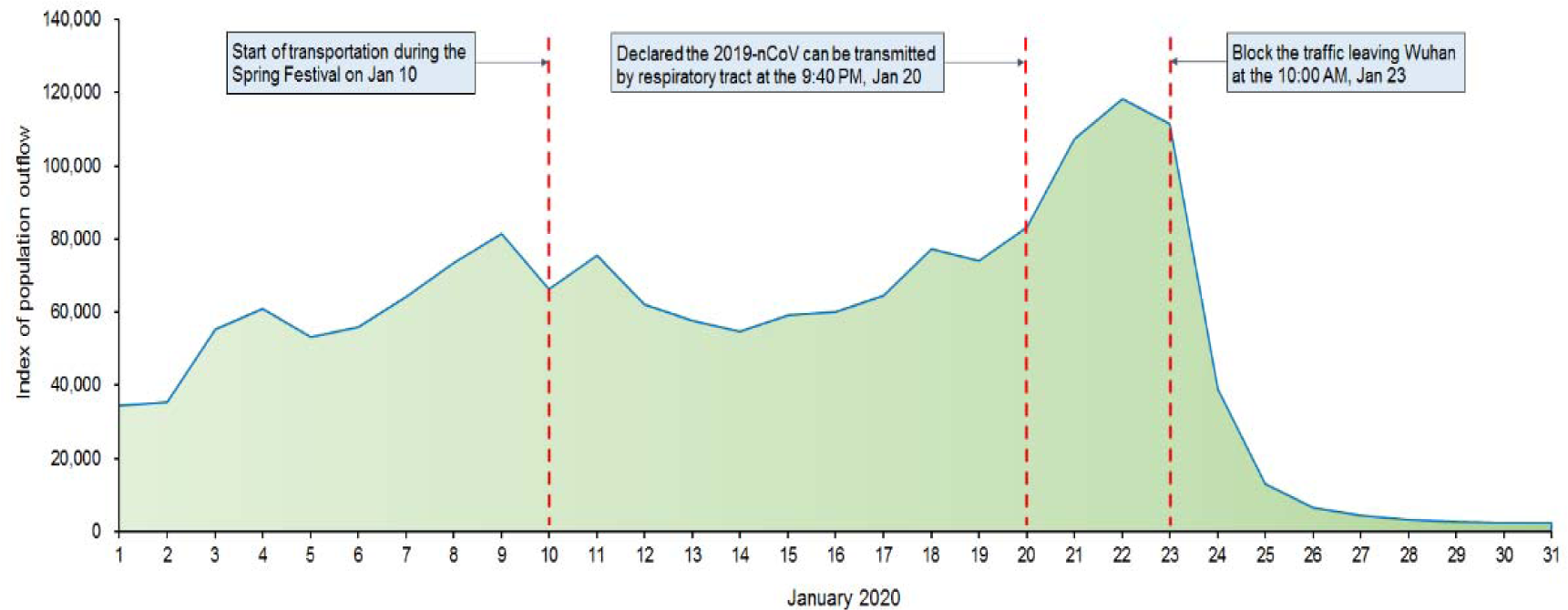
Index of population outflow of Wuhan City during period of January 1-31, 2020.

We applied scatter diagram to demonstrate the association between the number of 2019-nCoV cases and index of outflow population at the scales of province and city. In general, at the scale of province (figure 3), outflow population from Wuhan was mainly distributed in Henan, Hunan, Guangdong, Anhui, Zhejiang, etc. And the top four provinces with the most serious epidemic were Zhejiang, Guangdong, Henan, and Hunan provinces. Therefore, we could observe that there were more 2019-nCoV cases in the provinces with higher index of outflow population. Nevertheless, the two provinces of Zhejiang and Guangdong with relatively lower index of outflow population had more 2019-nCoV cases compared with Henan and Hunan provinces. In Henan, the index of outflow population was about 50000, while there were fewer than 450 cases, which might be lower than expected. In contrast, about 600 confirmed cases were observed in Zhejiang, which had only one fifth of outflow population of Henan. Similar results were obtained using the number cases per unit outflow population (appendix). The mean value of the number of 2019-nCoV cases per unit outflow population across the whole country was 129·72, however, the value in Zhejiang province was more than 450 and the corresponding result in Henan was lower than average.

**Figure 3.**
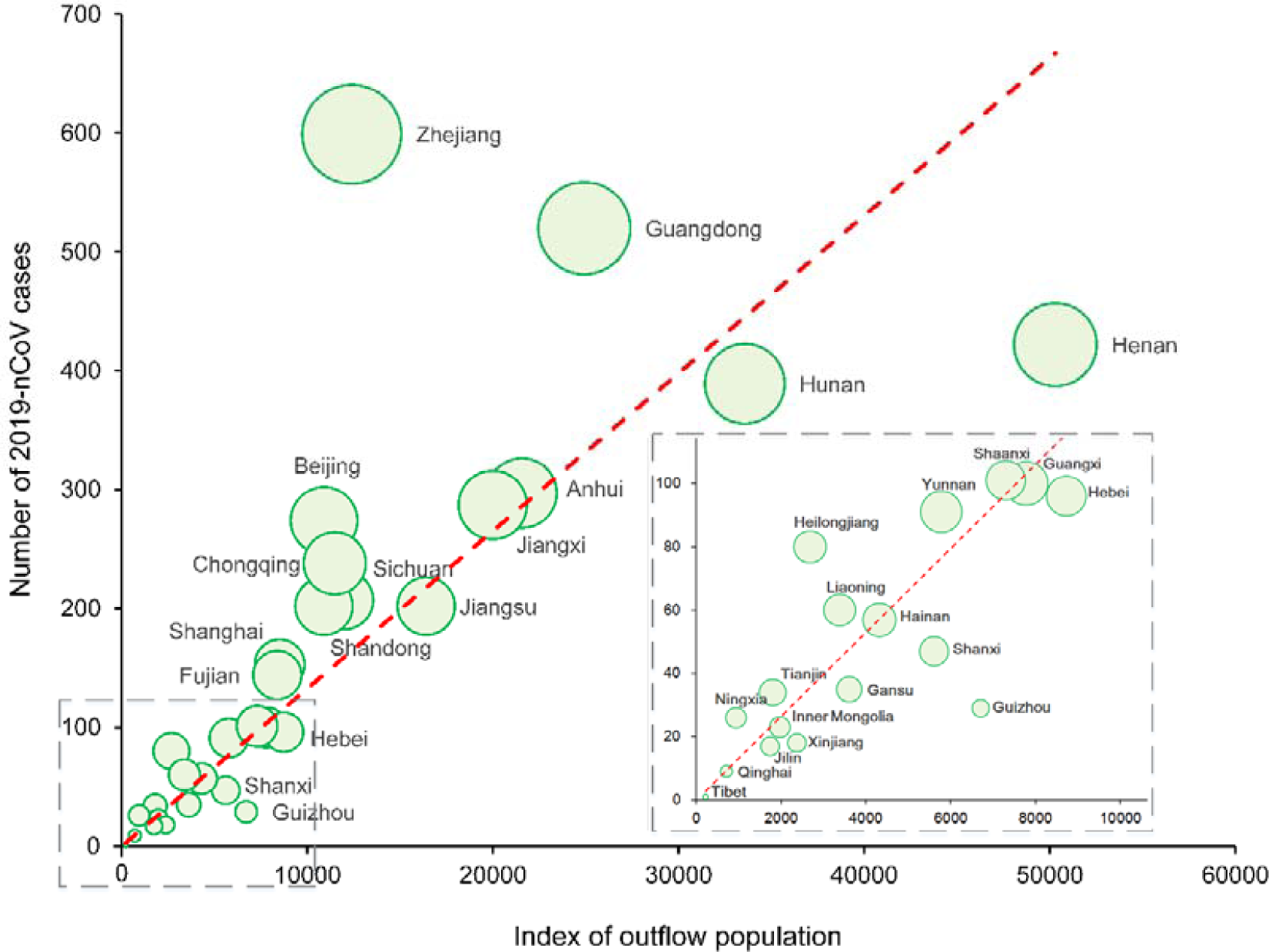
The association between the number of 2019-nCoV cases and the total index of population outflow at the provincial scale.

At the city-level analysis (figure 4), we observed that there was a large number of cases in the cities of Hubei province such as Huanggang, Xiaogan, Xiangyang, Jingzhou, etc. Overall, the association between the number of 2019-nCoV cases and index of outflow population at the city-level was in line with that at the provincial level, which both showed that the cities with more index of outflow population would trigger more cases. Similarly, we observed some cities deviated from the general trend including Wenzhou, Taizhou, Xuchang, Luoyang, and Guiyang. For instance, the index of outflow population of Wenzhou and Luoyang was close to identical level, whereas there was tremendous difference in the number of 2019-nCoV cases in the two cities. From the city of Taizhou and Xuchang, we could obtain the analogous results.

**Figure 4.**
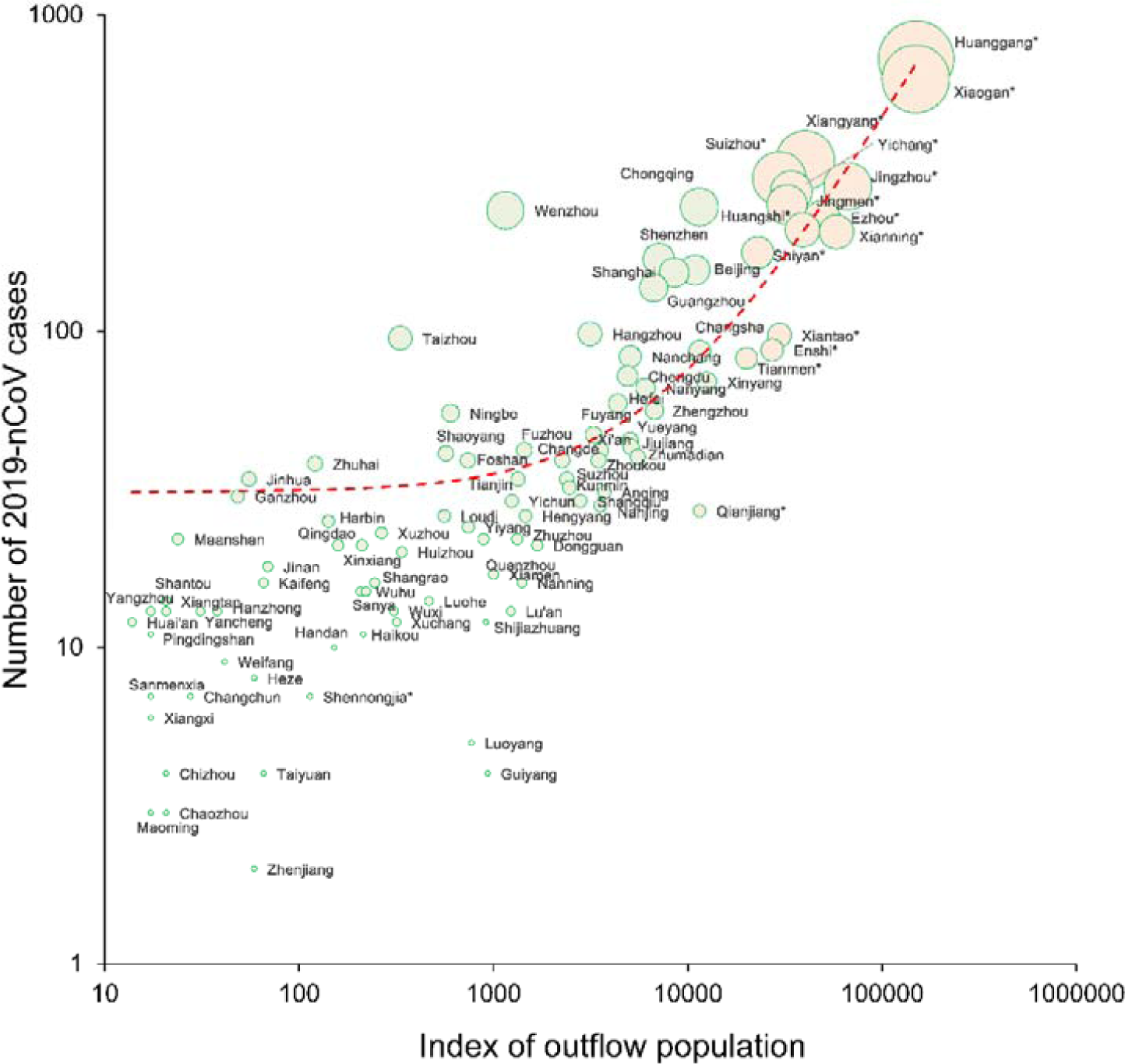
The association between the number of cases and the total index of population outflow at the city scale.

To evaluate the effect of the city closure on the infection transmission one and two days in advance, we used the index of population outflow of January 21-23, 2020 when the Wuhan was still open and index of population outflow of January 24-31, 2020 when the city closure policy had been implemented, and the calculations gave the reduced index of population outflow: 102206·69 for city closure one day in advance and 211429·60 for city closure two days in advance. In addition, we obtained the reduced index of population outflow of each other province and correspondingly reduced 2019-nCoV cases. Finally, about 687 and 1420 2019-nCoV cases would be avoided with implementing city closure policy one and two days in advance (table 1). Consistently, if city closure policy of Wuhan was delayed by one or two days, there would be 722 or 1462 extra 2019-nCoV cases respectively.

**Table 1.** The impact of different city closure dates on the number of 2019-nCoV infections in China.

| Alternative implementing dates | Reduced or increased 2019-nCoV cases* | 95% CI* |
| --- | --- | --- |
| Two days earlier: 2020-01-21 | -1420 | -1833, -1059 |
| One day earlier: 2020-01-22 | -687 | -886, -512 |
| One day later: 2020-01-24 | 722 | 539, 932 |
| Two days later: 2020-01-25 | 1462 | 1090, 1886 |

\*Table footnotes: 2019-nCoV=2019 novel coronavirus. CI=confidence interval.

## Discussion

Understanding the driving factors of the infectious disease is of particular importance for the timely formulation of effective controlling measures. Our results revealed a close relationship between the population outflow from Wuhan and the 2019-nCoV infection in other areas of China. We assessed the contributions of the lockdown and explored the potential effects of the measure with different implementation dates.

The fluctuation of outflowing population of Wuhan indicated that the lockdown in Wuhan reduced the outward movement of the population effectively. However, the migration data also suggested that a large number of populations of Wuhan had flowed out before 10:00 am Jan 23, 2020. and 2019-nCoV cases has been widely dispersed nationwide.

Distribution of population outflows and cases varied across provinces. Our finding on the effects of population movement on the disease transmission was consistent with other coronaviruses (16-18). Since the infection is transmitted through the respiratory tract and close contact, it is greatly affected by population movements. Firstly, a great number of people flowing out of Wuhan may transmit the virus to the whole country and even worldwide during the Chinese Spring Festival. Secondly, the absence of detectable symptoms during the long incubation period made it difficult to identify the cases in the early stage (13, 19). Thirdly, population movements increase the difficulty of case management and health education. All these factors promoted the transmission of 2019-nCoV. Specially, different from the H1N1 outbreak in 2009 (20), the railways might play an important role in the disease transmission. Since railways were widely used before and during the Spring Festival, the area closely connected to Wuhan by railway should be the priority of infection control.

Our study provided timely evidence for the formulation of efficient strategies to prevent diseases from spreading out. On the one hand, the result could help assess effectiveness of the prevention and control efforts. For example, the cases in Zhejiang and Guangdong are apparently more than estimated, which indicated a better health emergency response system (i.e. higher detection efficiency) or inadequate isolation. Whereas the cases reported in Henan were much lower than expected. Two possible explanations were considered: (1) Strong prevention and control measures were adopted in Henan; (2) The epidemic in Henan has been underestimated and enhanced screening efforts should be enforced. On the other hand, exploring the association was expected to help identify high-risk areas and guide health strategy formulation (21, 22). Take Henan for example, great difference between estimated and reported data may imply a great increase of cases in the future, which required enhancement of the surveillance system and rational allocation of resources (18). The medicine supply, personal protective equipment, hospital supplies, and the human resources necessary to respond to an outbreak should be always ensured (23). In addition, this study could be used to guide the assessment of the risk of disease transmission and help raise public awareness. As a large number of infected people had transported to all of 31 provinces, epidemics across the country may be inevitable. To halt the spread of the epidemic, harsh measures including quarantine and isolation of exposed persons, cancellation of mass gatherings, school closures, and travel restriction were needed to reduce transmission in affected areas. Furthermore, screening of people who have been to Wuhan recently was of crucial importance, especially cities with close ties to Wuhan.

Considering the impact of population movements on the outbreak, the Wuhan government announced the suspension of public transportation on Jan 23, 2020, with a closure of airports, railway stations, and highways, to prevent further disease transmission (24). Despite inconsistent reports on the role of the lockdown in halting the disease transmission across China (22, 23, 25), the unprecedented measure might play an important role in slowing the epidemic spread, especially when an effective vaccine was developed (26, 27). In addition, to explore the impact of date selection, we estimated the changes of cases when the measure was implemented on different date. The results varied significantly, 1420 cases could be prevented with the measure implemented two days earlier, and the number of cases will increase by 1462 with the lockdown implemented two days later, suggesting that the effect of the lockdown depending on the choice of date greatly, which could provide a reference for the future outbreaks. Since the political and economic effects were not considered, further studies on secondary impacts of the measure, like socioeconomic impacts, were also warranted. Though we estimated that some cases would possibly be prevented if the policy was implemented earlier, it was actually hard to make such a huge decision given the whole picture of the infection was not clear at that stage. The authors believe that the current policy was appropriate at this complex situation.

There were a few limitations of our study. Firstly, for practical reasons, we used an indicator to reflect the real-time magnitude of population movements, which was acceptable considering our research purpose. Secondly, the influence of some important factors, such as socioeconomic and demographic characteristics, were not considered. Thirdly, it is assumed that the infected travelers in the population are randomly distributed (28), and that there was no significant difference in surveillance capability between cities (18), which might not be the case in reality. In addition, daily data used in this study was reported infection data, rather than the actual number of incident cases. More detailed and further in-depth studies are warranted in the future.

In conclusion, our study indicates that the population outflow from Wuhan might be one important trigger of the 2019-nCoV infection transmission in China, and the policy of city closure is effective to prevent the epidemic and earlier implementation would be more effective. The magnitude of epidemic might be under-estimated and should be paid more attention, such as Henan and Hunan provinces.

## Data Availability

All URLs of the database related to the manuscript are provided as follows.

http://www.nhc.gov.cn/

http://qianxi.baidu.com

## Acknowledgments

We appreciated the support by National Key R&D Program of China (Grant No: 2018YFA0606200). The funder had no role in study design, data collection, data analysis, data interpretation, or writing of the report. The corresponding author had full access to all the data in the study and had final responsibility for the decision to submit for publication.

## About the Author

Mr. Ai completed this work as a Ph.D. student in Department of Epidemiology, School of Public Health, Sun Yat-sen University, his primary research direction is the spatio-temporal impact of climate change and air pollution on human health.

